# Exhaled SARS-CoV-2 quantified by face-mask sampling in hospitalised patients with covid-19

**DOI:** 10.1101/2020.08.18.20176693

**Authors:** Caroline M. Williams, Daniel Pan, Jonathan Decker, Eve Fletcher, Anika Wisniewska, Shirley Sze, Sara Assadi, Richard Haigh, Mohamad Abdulwhhab, Paul Bird, Christopher W Holmes, Alaa Al-Taie, Baber Saleem, Jingzhe Pan, Natalie J Garton, Manish Pareek, Michael R Barer

**Author notes:** Joint first authors (Caroline M. Williams, Daniel Pan, Jonathan Decker). Join senior authors (Manish Pareek, Michael R Barer). **Corresponding authors**: Dr Caroline Williams, Dr Manish Pareek, Professor Mike Barer.

## Abstract

**Background:** Human to human transmission of SARS-CoV-2 is driven by the respiratory route but little is known about the pattern and quantity of virus output from exhaled breath. We have previously shown that face-mask sampling (FMS) can detect exhaled tubercle bacilli and have adapted its use to quantify exhaled SARS-CoV-2 RNA in patients admitted to hospital with covid-19.

**Methods:** Between May and December 2020, we took two concomitant FMS and nasopharyngeal samples (NPS) over two days, starting within 24 hours of a routine virus positive NPS in patients hospitalised with covid-19, at University Hospitals of Leicester NHS Trust, UK. Participants were asked to wear a modified duckbilled facemask for 30 minutes, followed by a nasopharyngeal swab. Demographic, clinical, and radiological data, as well as International Severe Acute Respiratory and emerging Infections Consortium (ISARIC) mortality and deterioration scores were obtained.

Exposed masks were processed by removal, dissolution and analysis of sampling matrix strips fixed within the mask by RT-qPCR. Viral genome copy numbers were determined and results classified as Negative; Low: ≤999 copies; Medium: 1,000-99,999 copies and High ≥ 100,000 copies per strip for FMS or per 100µl for NPS.

**Results:** 102 FMS and NPS were collected from 66 routinely positive patients; median age: 61 (IQR 49 - 77), of which FMS was positive in 37% of individuals and concomitant NPS was positive in 50%. Positive FMS viral loads varied over five orders of magnitude (<10-3.3 × 10^6^ genome copies/strip); 21 (32%) patients were asymptomatic at the time of sampling. High FMS viral load was associated with respiratory symptoms at time of sampling and shorter interval between sampling and symptom onset (FMS High: median (IQR) 2 days (2-3) vs FMS Negative: 7 days (7-10), *p*=0.002). On multivariable linear regression analysis, higher FMS viral loads were associated with higher ISARIC mortality (Medium FMS vs Negative FMS gave an adjusted coefficient of 15.7, 95% CI 3.7-27.7, *p*=0.01) and deterioration scores (High FMS vs Negative FMS gave an adjusted coefficient of 37.6, 95% CI 14.0 to 61.3, *p*=0.002), while NPS viral loads showed no significant association.

**Conclusion:** We demonstrate a simple and effective method for detecting and quantifying exhaled SARS-CoV-2 in hospitalised patients with covid-19. Higher FMS viral loads were more likely to be associated with developing severe disease compared to NPS viral loads. Similar to NPS, FMS viral load was highest in early disease and in those with active respiratory symptoms, highlighting the potential role of FMS in understanding infectivity.

## INTRODUCTION

Severe acute respiratory syndrome–coronavirus-2 (SARS-CoV-2) is predominantly transmitted between humans by the respiratory route.^1^ While nasopharyngeal sampling (NPS) is the current standard method for the diagnosis of covid-19, its role in identification of infectious individuals is limited.^2^ There is emerging evidence to suggest that virus detected in the exhaled breath of infected patients significantly contribute to SARS-CoV-2 transmission.^3–5^ However, little is known about the quantity and pattern of output.

The use of face-masks in an infected individual significantly reduces detection of influenza virus RNA in respiratory droplets and seasonal coronavirus RNA in aerosols.^6^ Therefore, collection of respiratory samples in face-masks where emitted pathogen densities are highest^7–9^, offers potential advantages. We have been developing this approach, which we term face-mask sampling (FMS), to detect and quantify exhaled pathogens, initially to study tuberculosis.^10–12^ In this study, we describe the use of FMS in hospitalised patients with covid-19. We investigated relationships between viral load in FMS and NPS and determined associations with disease severity in patients at different stages of acute infection.

## METHODS

### Study setting

We undertook a prospective observational study which enrolled patients hospitalised for covid-19 at University Hospitals of Leicester NHS Trust, Leicester UK. Up to two serial FMS and NPS samples were taken for analyses, with the first samples taken within 24 hours of a routinely positive NPS test. We included patients who fulfilled the following criteria: age ≥16 years, hospitalised, tested positive for SARS-CoV-2 on routine NPS testing (D0) and no requirement for oxygen therapy by face-mask, non-invasive ventilation or intubation (since this would prevent using face-masks from the study) at time of sampling. Patients who were unable to understand and comply with the protocol, or unable or unwilling to give informed consent, were not included in the study.

### Clinical data, ISARIC mortality and deterioration scores

We collected clinical data collected on: age, gender, ethnicity, smoking status, comorbidities (diabetes, hypertension, cardiac disease, chronic kidney disease, chronic lung disease, cerebrovascular disease, cancer and immunosuppression), hospital acquired covid-19, clinical symptoms at time of sampling, the duration between symptom onset and clinical observations at both time of sampling and at presentation with radiology and laboratory findings at time of presentation. Clinical outcomes collected included death within hospitalisation and rehospitalisation, reinfection or death by 21^st^ January 2021.

ISARIC mortality and deterioration scores were calculated. These scores are validated risk stratification tools that predict in-hospital mortality or in-hospital clinical deterioration (defined as any requirement for ventilator support or critical care, or death) for hospitalised covid-19 patients in the UK.^13,14^ The mortality score consists of: gender, number of comorbidities, Glasgow Coma score (GCS), age, respiratory rate, admission oxygen saturations, serum urea, C-reactive protein (CRP) and lymphocyte count. The deterioration score consists of: nosocomial covid-19, gender, number of comorbidities, radiographic chest infiltrates, whether the patient was receiving oxygen, GCS, age, respiratory rate, admission oxygen saturations, urea, CRP and lymphocyte count. For each, the raw score is converted into a calculated % risk of either in-hospital mortality or clinical deterioration.

### Sampling

Each participant wore a duckbilled surgical mask containing two or four 1×9 cm 3D printed polyvinyl-alcohol (PVA) sampling matrix strips placed horizontally across the front of the mask.^15^ Participants wore the mask for 30 minutes. No special behaviour was required, aside from not eating. Each participant was observed during the period in which they wore the mask to ensure that there were no periods where the mask was lifted. Following instruction, participants were sampled first within 24 hours (D1) of a routinely positive NPS test and a second time 12-24 hours later (D2); they were directly observed throughout FMS and the NPS was taken afterwards. Exposed masks and swabs were delivered to the laboratory and maintained at ambient temperature until analysis. In *in-vitro* preparatory studies, viral loads evaluated by RT-qPCR from PVA strips spiked with cultured SARS-CoV-2 and stored dry at room temperature were shown to be stable over seven days, while assay of unprocessed exposed strips similarly stored gave values comparable to those from the same mask processed within seven days of receipt and indicated stability over three months (supplementary materials).

The study had ethical approval from the West Midlands Research Ethics Committee (REC Reference 20/WM/0153) and was conducted in accordance with ICH-GCP, Declaration of Helsinki and the Data Protection Act 1998 and NHS Act 2006. All participants gave written, informed consent prior to any study procedures.

### Sample processing and controls

A detailed description of laboratory processing is provided in supplementary materials 2. In summary, for FMS processing, two PVA strips were dissolved in a mixture of molecular grade water and QIAamp ACL buffer and underwent RNA extraction using the QIAamp DSP Circulating Nucleic Acid Kit (Qiagen, Germany Cat 61504,) following the manufacturer’s instructions. For NPS, the sampled material was first eluted from the swab head into water by vortexing then RNA extracted using RNeasy mini kits (Qiagen, Cat 74104). For both sample types, target RNA was detected and quantified using the QuantiNova Probe RT-PCR Kit (Qiagen, Cat: 208356) and a Rotor-Gene Q thermocyler (Qiagen, Cat 9001590). Internal controls were added to every sample prior to extraction and a positive control sample included in every run; sample positivity/negativity was determined with assays directed to the E gene. All positive samples were quantified for genome copy number^16^ in a single E gene-directed PCR run for which the standard curve is shown in supplementary materials; for reference, 2,240 genome copies gave a Ct of 30. For *in vitro* studies, SARS-CoV-2 was isolated in Vero E6 cell culture by limiting dilution of a RT-qPCR positive swab sample; confirmation of isolate identity at passage 3 was undertaken by E assay.

### Statistical analysis

Continuous variables are expressed as median and interquartile range (IQR). Categorical variables are displayed as numbers and percentages (%). Pearson’s Chi-squared test and Fisher’s exact row test were used to compare categorical variables between groups. Student’s *t*-test and Kruskal-Wallis test were used to compare continuous variables between groups depending on the normality of distribution. We chose to examine the relation between increasing FMS and NPS viral load and clinical variables. The distribution of FMS and NPS viral loads obtained was log normally distributed (shown in supplementary materials 4) – therefore we classified the exhaled viral load as genome copies into the following four categories: Negative: PCR negative, Low: ≤999 copies, Medium:1000-99,999 copies and High: >100,000 copies per strip or per 100µl.

Multivariable linear regression was used to investigate the relationship between variables and increased risk of in-hospital mortality and deterioration, as measured by the validated ISARIC mortality and deterioration scores. Univariable linear regression was performed using clinical variables relating to increased disease severity, but not already included in the ISARIC scoring systems. Variables were chosen to be included in the models if they were significantly related to the relevant ISARIC score on univariable linear regression analysis (a threshold of *p*<0.05 was considered significant).

The agreement between FMS and NPS positivity was assessed using Cohen’s Kappa. Prediction of viral loads between FMS and NPS was assessed using linear regression. Scatter plots were used to illustrate the relation between FMS and NPS genome copies and time from symptom onset.

All analyses were performed using STATA version 14.2 (StataCorp, United States) and Excel version 2016 (Microsoft, Redmond, United States).

## RESULTS

### Participants

Between May and December 2020, 66 patients were enrolled. 36 patients provided two serial FMS and NPS samples; thus a total of 102 concomitant FMS and NPS samples were collected. For some patients, a second (D2) sample was not obtained for a number of reasons including: worsening of clinical disease (12, 18%), increased oxygen requirement (10, 15%), discharge from hospital (3, 5%) and reported discomfort while wearing the mask (5, 8%).

The median (IQR) age of participants was 61 (49-77). Twenty-four (60%) were male. Participants were predominantly of White ethnic background (47, 65%). Most patients (39, 59%) were sampled the day after their first known positive NPS PCR test; 18 (27%) were sampled after an interval of seven or more days. One third (21, 32%) of patients had acquired covid-19 in hospital and one third (21,32%) were asymptomatic at the time for the D1 FMS/NPS samples. For those who were symptomatic, the median duration of symptoms was seven (3-7) days prior to D1. No patient recruited had been vaccinated against covid-19, or taken part in any vaccination trials at the time of sampling.

Seventy-four (49%) patients had evidence of covid-19 pneumonia on their chest x-ray. The median serum urea was: 5.9 (4.2-8.8) mmol/L; CRP: 49 (15-110) mg/L and lymphocyte count: 1.13 (0.7-1.59) ×10^9^ /L. The median ISARIC mortality score (interquartile range, IQR) was 13% (5%-27%) and median deterioration score was 28% (15%-43%). Five patients died during hospital admission; a further five died after discharge from hospital (10 patients),

### Detection of viral RNA on FMS and comparison with NPS

RNA recovered from the PVA sampling strip matrix used in FMS was positive in 41/102 samples (40%) from 25/66 (37%) of individuals. NPS was positive in 51/102 (50%) samples from 33/66 (50%) of individuals. In patients whose concomitant NPS was positive, D1 FMS was positive in 20/31 (65%) individuals while the D2 FMS was positive in 14/20 (70%) of individuals. There was a moderate agreement between concomitant positive FMS and NPS samples (Cohen’s kappa: 0.52 *p*<0.001,) and the log_10_ viral loads were weakly, though significantly, correlated (r^2^=0.15, *p*=0.022) as shown in Figure 1. Amongst those that were both FMS and NPS positive, there were five instances where participants had a disproportionately higher FMS viral load than predicted by NPS viral load (red symbols in Figure 1). In total, 37 (56%) patients tested positive by either FMS or NPS. Five patients were FMS positive NPS negative (viral loads in all of these samples were in the Low category). Twelve patients were NPS positive and FMS negative (viral loads in Low, Medium and High categories).

**Figure 1.**
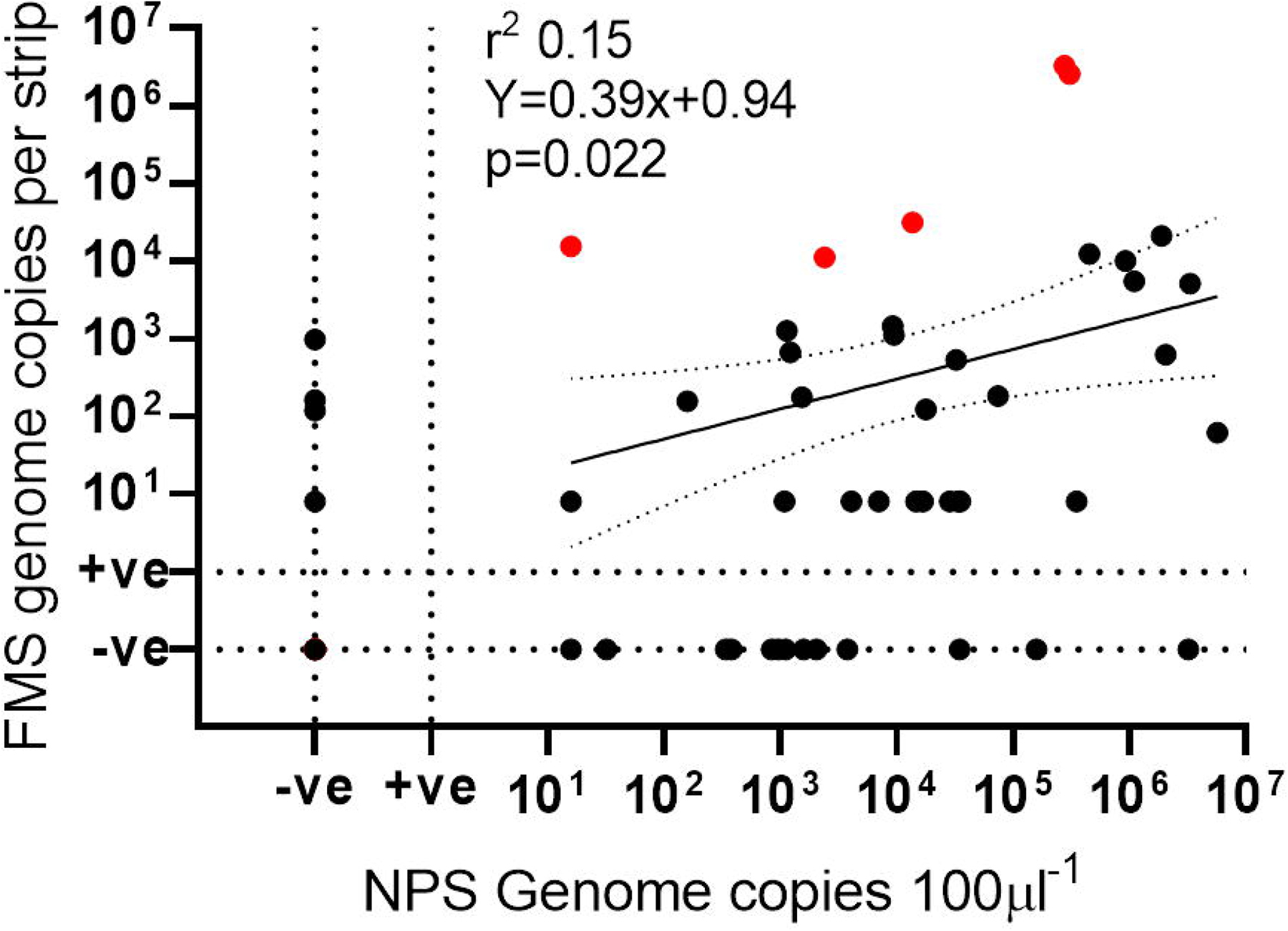
Relationship between FMS and NPS viral loads. Abbreviations used: FMS – face-mask sampling; NPS – nasopharyngeal sampling. Red data points represent those with a disproportionately higher FMS viral load than predicted by NPS viral load.

### Relation of FMS to clinical variables

FMS positive E target genome copy numbers ranged from 8 to 3.3 × 10^6^ per strip (values assigned to borderline positives). Ten patients (15%) had a positive FMS sample in the Medium viral load range (10^3^-10^5^ copies/strip); High loads were seen in two patients (3%) in their second sample (2.6 and 3.3 × 10^6^ copies/strip).

Patients with higher FMS viral load were more likely to be older (*p*=0.04) and to have acquired covid-19 in hospital (*p*=0.004), as shown in Table 1. They were also more likely to have elevated serum urea on admission (*p*=0.03). NPS samples showed twelve (18%) patients with viral loads of 10^3^-10^6^ copies/100µl swab suspension and nine (14%) with loads between 10^5^ and 10^7^ copies/100µl. Higher viral loads on FMS or NPS were found in patients who had active respiratory symptoms at time of sampling (cough: FMS *p*=0.02; NPS *p*=0.01; breathlessness: FMS *p*=0.01; NPS *p*=0.01) and with shorter intervals between sampling and day of symptom onset (FMS *p*=0.007; NPS *p=* 0.002) as shown in Table 1 and Figure 2. There was no relation between FMS or NPS viral load to fever or other symptoms (arthralgia, fatigue and anosmia), or evidence of covid-19 pneumonia on chest x-ray.

**Table 1:** Demographics of participants in relation to increasing FMS and NPS genome copy numbers. Abbreviations used: FMS – face-mask sampling; NPS – nasopharyngeal sampling

| Variables (n=66) | FMS genome copy numbers (copies/strip) |  |  |  |  | NPS genome copy numbers (copies/100 microlitres) |  |  |  |  |
| --- | --- | --- | --- | --- | --- | --- | --- | --- | --- | --- |
|  | Negative n=40 | Low n=14 | Medium n=10 | High n=2 | P value | Negative n=33 | Low n=12 | Medium n=12 | High n=9 | P value |
| Demographics |  |  |  |  |  |  |  |  |  |  |
| Age – years | 58 (42-73) | 68 (45-84) | 71 (62-79) | 79 (69-89) | 0.04 | 58 (42-71) | 69 (55-81) | 61 (47-77) | 71 (63-80) | 0.21 |
| Males – (%) | 24 (60%) | 8 (57%) | 9 (90%) | 2 (100%) | 0.24 | 20 (61%) | 7 (58%) | 9 (75%) | 7 (78%) | 0.72 |
| Ethnicity |  |  |  |  |  |  |  |  |  |  |
| White – (%) | 28 (70%) | 10 (72%) | 7 (70%) | 2 (100%) | 0.54 | 22 (70%) | 10 (83%) | 8 (67%) | 6 (67%) | 0.71 |
| Asian – (%) | 11 (28%) | 2 (14%) | 3 (30%) | 0 (0%) |  | 8 (24%) | 1 (9%) | 4 (33%) | 3 (34%) |  |
| Black – (%) | 1 (3%) | 2 (14%) | 0 (0%) | 0 (0%) |  | 2 (6%) | 1 (8%) | 0 (0%) | 0 (0%) |  |
| Smoker |  |  |  |  |  |  |  |  |  |  |
| Ever smoked– (%) | 27 (68%) | 10 (71%) | 9 (90%) | 2 (100%) | 0.93 | 26 (81%) | 7 (58%) | 9 (75%) | 6 (67%) | 0.34 |
| Never – (%) | 10 (25%) | 3 (22%) | 1 (10%) | 0 (0%) |  | 6 (18%) | 4 (33%) | 1 (8%) | 3 (33%) |  |
| Nosocomial covid-19 – (%) | 6 (15%) | 7 (50%) | 4 (40%) | 2 (100%) | 0.004 | 6 (18%) | 6 (50%) | 4 (33%) | 3 (33%) | 0.25 |
| Comorbidities |  |  |  |  |  |  |  |  |  |  |
| <2 – (%) | 25 (62%) | 5 (36%) | 3 (30%) | 2 (100%) | 0.06 | 20 (61%) | 5 (42%) | 7 (58%) | 1 (11%) | 0.06 |
| ≥2 – (%) | 15 (38%) | 9 (64%) | 7 (70%) | 0 (0%) |  | 13 (39%) | 7 (58%) | 5 (42%) | 8 (89%) |  |
| Symptoms at time of sampling |  |  |  |  |  |  |  |  |  |  |
| Respiratory symptoms – (%) | 1 (45%) | 11 (79%) | 9 (90%) | 1 (50%) | 0.01 | 15 (46%) | 6 (50%) | 9 (75%) | 9 (100%) | 0.01 |
| Fever – (%) | 21 (53%) | 8 (57%) | 5 (50%) | 1 (50%) | 0.99 | 15 (46%) | 7 (58%) | 6 (50%) | 7 (78%) | 0.40 |
| Other – (%) | 16 (40%) | 8 (57%) | 6 (60%) | 1 (50%) | 0.54 | 12 (36%) | 5 (42%) | 7 (58%) | 7 (78%) | 0.14 |
| Asymptomatic – (%) | 16 (40%) | 3 (21%) | 1 (10%) | 1 (10%) | 0.19 | 13 (39%) | 5 (42%) | 3 (25%) | 0 (0%) | 0.10 |
| Symptom duration – (%) | 7 (6-10) | 5 (2-7) | 3 (3-3) | 1 (1-1) | 0.007 | 7 (7-10) | 7 (2-8) | 7 (2-7) | 3 (2-3) | 0.002 |
| Radiological and laboratory investigations |  |  |  |  |  |  |  |  |  |  |
| Features of Covid-19 on chest x-ray – (%) | 28 (70%) | 10 (71%) | 9 (90%) | 2 (100%) | 0.60 | 23 (70%) | 9 (75%) | 9 (75%) | 8 (89%) | 0.80 |
| Receiving oxygen when measured – (%) | 9 (23%) | 1 (7%) | 3 (30%) | 0 (0%) | 0.48 | 6 (18%) | 4 (33%) | 2 (7%) | 1 (11%) | 0.66 |
| Outcomes |  |  |  |  |  |  |  |  |  |  |
| Died – (%) | 6 (15%) | 1 (7%) | 3 (30%) | 0 (0%) | 0.46 | 10 (30%) | 5 (42%) | 3 (25%) | 7 (78%) | 0.06 |
| ISARIC mortality score – (% risk of hospital death) | 12 (2-21) | 12 (7-23) | 37 (19-45) | 41 (23-59) | 0.006 | 12 (2-23) | 25 (7-37) | 12 (2-21) | 23 (12-40) | 0.04 |
| ISARIC deterioration score – (% risk of hospital deterioration) | 28 (16-41) | 18 (14-31) | 49 (28-60) | 62 (42-82) | 0.01 | 27 (15-39) | 39 (16-55) | 27 (13-33) | 31 (22-44) | 0.63 |

**Figure 2.**
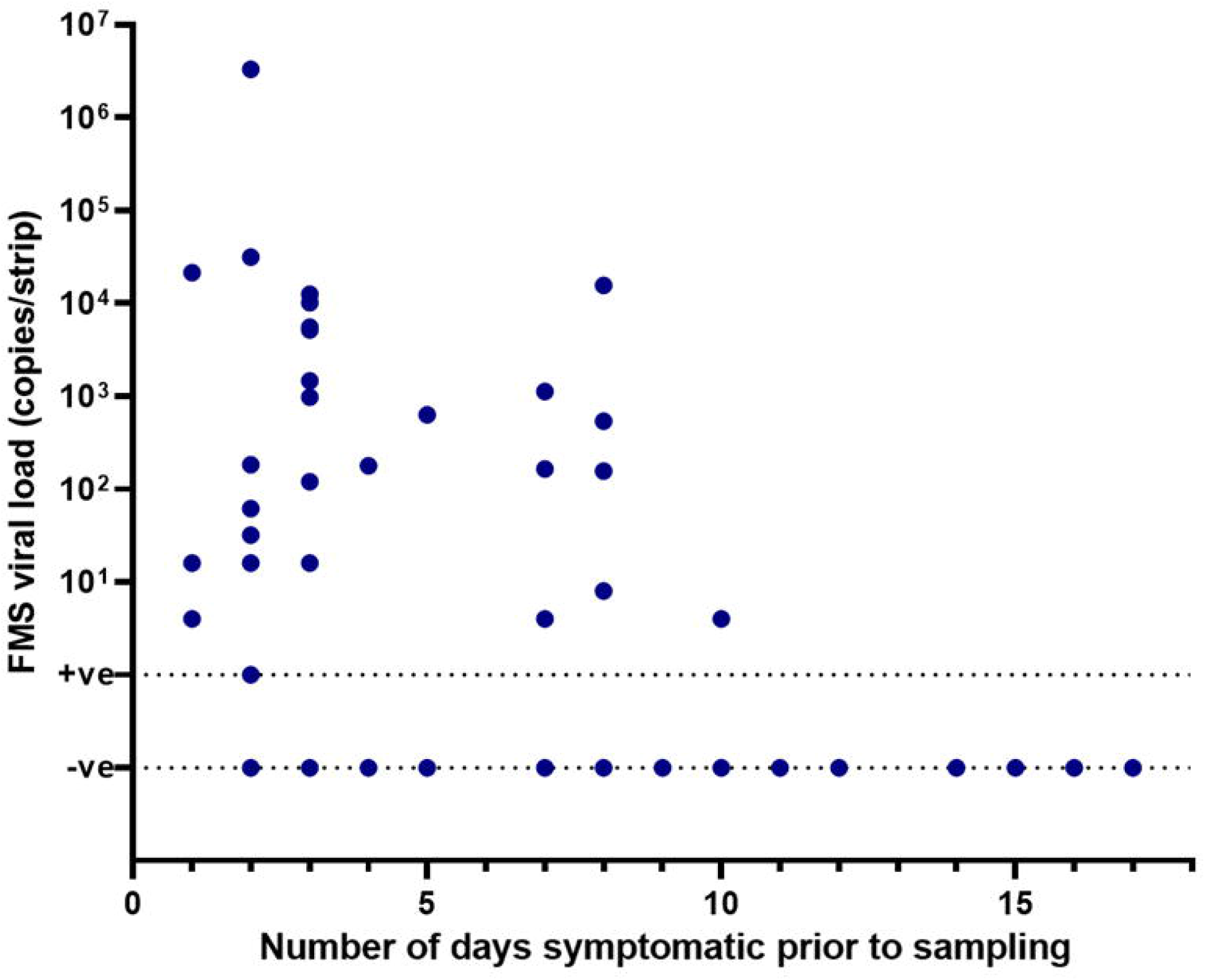
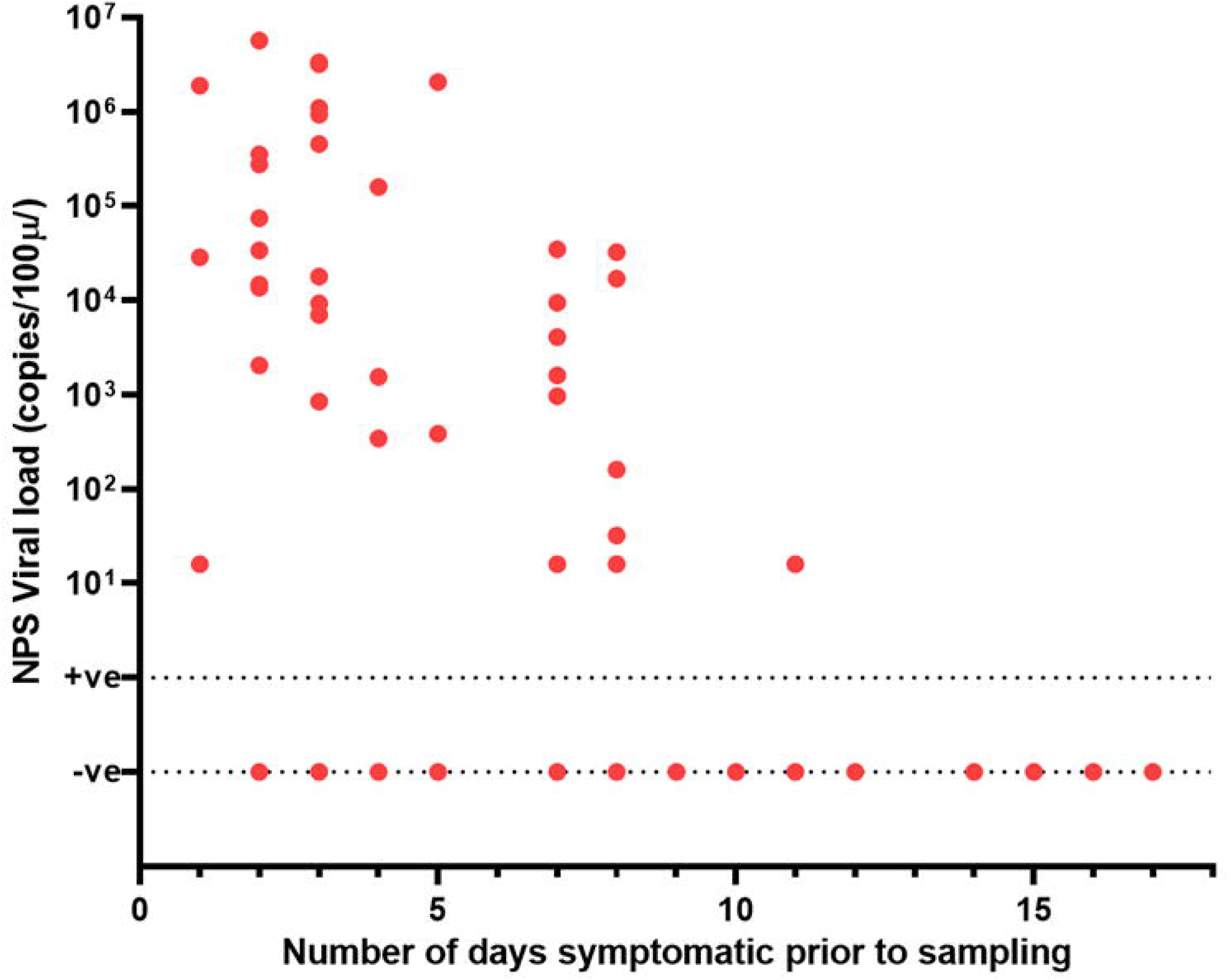
FMS (2a) and NPS (2b) viral copy numbers in relation to number of days symptomatic prior to sampling. Abbreviations used: FMS – face-mask sampling; NPS – nasopharyngeal sampling

On univariable linear regression analysis, nosocomial infection (*p*=0.01), treatment with dexamethasone (*p*=0.01), infiltrates on chest x-ray (*p*=0.02) and increasing FMS and NPS viral loads (FMS: *p*<0.001; NPS: *p*=0.02) were associated with higher ISARIC mortality scores (Table 2). Those from Asian minority groups had lower ISARIC mortality scores compared to White groups (*p*=0.01). On multivariable analysis, higher ISARIC mortality scores were found in those with higher FMS viral loads (*p*=0.01), as well as those of White ethnicity compared to Asian groups (*p*=0.02). Treatment with dexamethasone and a High FMS viral load was associated with higher ISARIC deterioration scores on both univariable and multivariable linear regression analysis (multivariable analysis, dexamethasone: *p*<0.001, FMS viral load: *p*=0.002).

**Table 2:**
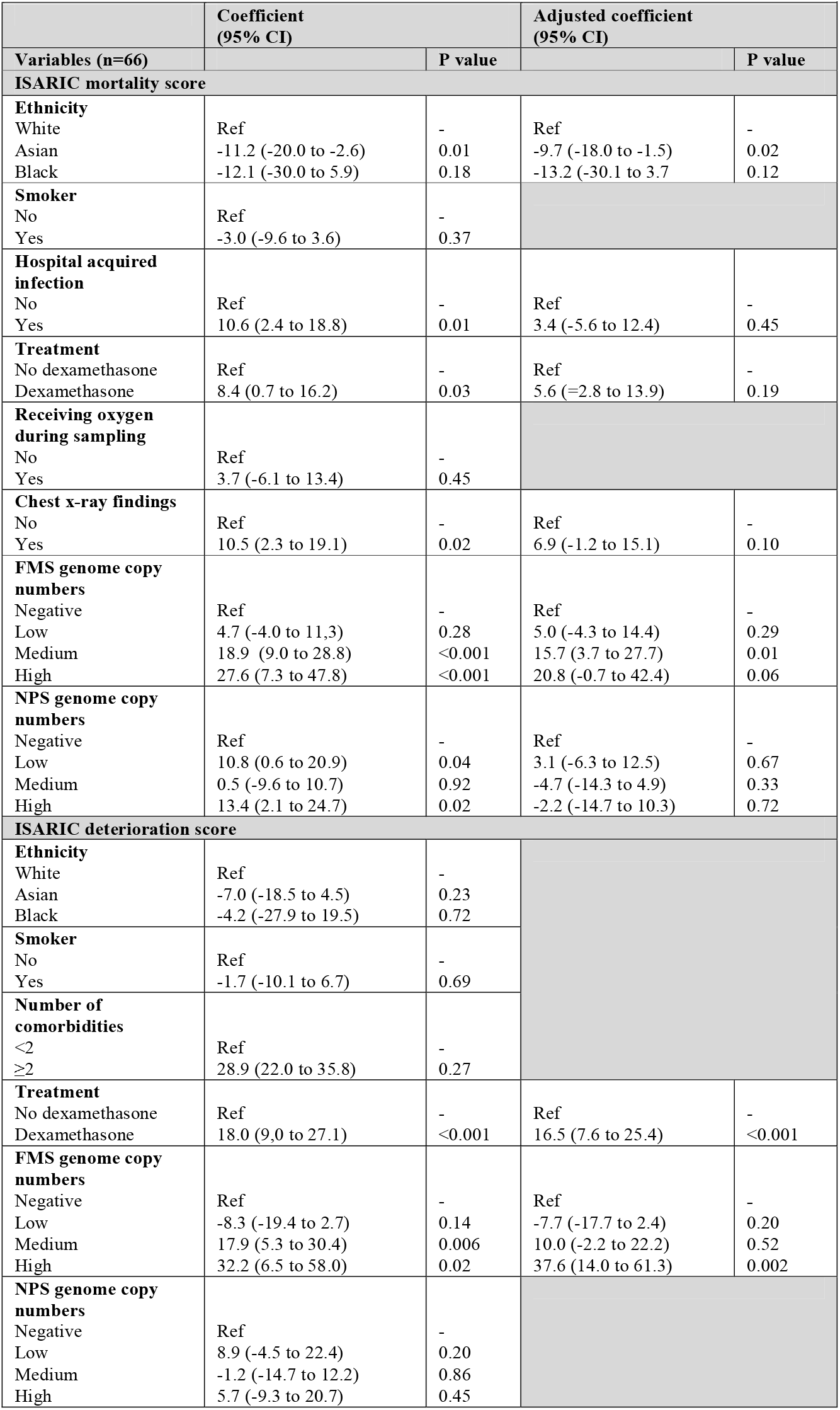
Univariable and multivariable linear regression models. Abbreviations used: FMS – face-mask sampling; NPS – nasopharyngeal sampling

|  | Coefficient<br>(95% CI) |  | Adjusted coefficient<br>(95% CI) |  |
| --- | --- | --- | --- | --- |
| Variables (n=66) |  | P value |  | P value |
| **ISARIC mortality score** |  |  |  |  |
| **Ethnicity** |  |  |  |  |
| White | Ref | - | Ref | - |
| Asian | -11.2 (-20.0 to -2.6) | 0.01 | -9.7 (-18.0 to -1.5) | 0.02 |
| Black | -12.1 (-30.0 to 5.9) | 0.18 | -13.2 (-30.1 to 3.7) | 0.12 |
| **Smoker** |  |  |  |  |
| No | Ref | - |  |  |
| Yes | -3.0 (-9.6 to 3.6) | 0.37 |  |  |
| **Hospital acquired infection** |  |  |  |  |
| No | Ref | - | Ref | - |
| Yes | 10.6 (2.4 to 18.8) | 0.01 | 3.4 (-5.6 to 12.4) | 0.45 |
| **Treatment** |  |  |  |  |
| No dexamethasone | Ref | - | Ref | - |
| Dexamethasone | 8.4 (0.7 to 16.2) | 0.03 | 5.6 (=2.8 to 13.9) | 0.19 |
| **Receiving oxygen during sampling** |  |  |  |  |
| No | Ref | - |  |  |
| Yes | 3.7 (-6.1 to 13.4) | 0.45 |  |  |
| **Chest x-ray findings** |  |  |  |  |
| No | Ref | - | Ref | - |
| Yes | 10.5 (2.3 to 19.1) | 0.02 | 6.9 (-1.2 to 15.1) | 0.10 |
| **FMS genome copy numbers** |  |  |  |  |
| Negative | Ref | - | Ref | - |
| Low | 4.7 (-4.0 to 11.3) | 0.28 | 5.0 (-4.3 to 14.4) | 0.29 |
| Medium | 18.9 (9.0 to 28.8) | <0.001 | 15.7 (3.7 to 27.7) | 0.01 |
| High | 27.6 (7.3 to 47.8) | <0.001 | 20.8 (-0.7 to 42.4) | 0.06 |
| **NPS genome copy numbers** |  |  |  |  |
| Negative | Ref | - | Ref | - |
| Low | 10.8 (0.6 to 20.9) | 0.04 | 3.1 (-6.3 to 12.5) | 0.67 |
| Medium | 0.5 (-9.6 to 10.7) | 0.92 | -4.7 (-14.3 to 4.9) | 0.33 |
| High | 13.4 (2.1 to 24.7) | 0.02 | -2.2 (-14.7 to 10.3) | 0.72 |
| **ISARIC deterioration score** |  |  |  |  |
| **Ethnicity** |  |  |  |  |
| White | Ref | - |  |  |
| Asian | -7.0 (-18.5 to 4.5) | 0.23 |  |  |
| Black | -4.2 (-27.9 to 19.5) | 0.72 |  |  |
| **Smoker** |  |  |  |  |
| No | Ref | - |  |  |
| Yes | -1.7 (-10.1 to 6.7) | 0.69 |  |  |
| **Number of comorbidities** |  |  |  |  |
| <2 | Ref | - |  |  |
| ≥2 | 28.9 (22.0 to 35.8) | 0.27 |  |  |
| **Treatment** |  |  |  |  |
| No dexamethasone | Ref | - | Ref | - |
| Dexamethasone | 18.0 (9.0 to 27.1) | <0.001 | 16.5 (7.6 to 25.4) | <0.001 |
| **FMS genome copy numbers** |  |  |  |  |
| Negative | Ref | - | Ref | - |
| Low | -8.3 (-19.4 to 2.7) | 0.14 | -7.7 (-17.7 to 2.4) | 0.20 |
| Medium | 17.9 (5.3 to 30.4) | 0.006 | 10.0 (-2.2 to 22.2) | 0.52 |
| High | 32.2 (6.5 to 58.0) | 0.02 | 37.6 (14.0 to 61.3) | 0.002 |
| **NPS genome copy numbers** |  |  |  |  |
| Negative | Ref | - |  |  |
| Low | 8.9 (-4.5 to 22.4) | 0.20 |  |  |
| Medium | -1.2 (-14.7 to 12.2) | 0.86 |  |  |
| High | 5.7 (-9.3 to 20.7) | 0.45 |  |  |

## DISCUSSION

With this study, using methods applicable in routine clinical and laboratory settings, we have demonstrated detection and quantification SARS-CoV-2 genomes in the exhaled breath of infected patients, one third of whom were asymptomatic at time of sampling. We found that patients exhaled viral RNA ranging over five orders in magnitude and that this appeared to be highest in the first few days of symptoms and when active respiratory symptoms were present at time of sampling. In contrast to concomitant NPS viral loads, we also found that higher FMS viral loads may have an association with more severe disease.

FMS was able to detect SARS-CoV-2 in the exhaled breath of 37% of individuals that are known to have covid-19. This contrasts with the 26.9% positive rate in infected individuals, reported by Ma and colleagues who sampled for five minutes using a breath condensate sampling device.^5^ We note the use of a bespoke sampling device in the Ma study and the application of processing methods largely outside those generally available in routine settings. Our results are similar to the preliminary report of Sriraman and colleagues, who found 40% positivity rate amongst infected individuals by FMS using a gelatine sampling matrix.^17^

Previous studies have shown that NPS signals can be persistently high for several weeks despite index cases that are no longer infectious.^18–20^ Kim and colleagues found that in hospitalised patients with covid-19, the median time from symptom onset to viral clearance was 7 days.^21^ In our study, no patient was FMS positive for more than 10 days after symptom onset. FMS viral load was highest in early disease (including in those who were nosocomial infected) and more virus was exhaled in those with active respiratory symptoms at time of sampling than those who were asymptomatic.

The quantities of exhaled virus detected were not always predicted by quantitation by NPS. Linear regression analysis indicated that the NPS-detected viral RNA only accounts for 15% of the variation detected by FMS (r^2^ = 0.15). In particular, the detection of five individuals whose FMS viral loads greatly exceeded those predicted by the general relationship between FMS and NPS. FMS viral loads were low when NPS was negative; NPS viral load on the other hand varied significantly between samples when concomitant FMS was negative.

Altogether, these findings suggest that FMS viral load patterns emitted from individuals may be distinct from NPS. Future work will explore the relationship between patterns of FMS output of SARS-CoV-2 and transmission to determine the value of this approach in determining infectivity of individuals, compared to other tests proposed for similar purposes, such as the lateral flow assays.^22^

In multivariable analyses, higher FMS viral loads, but not those detected by NPS, were significantly related to both ISARIC mortality and deterioration scores. Previous studies also show little differences in NPS viral load between those with mild, moderate and severe disease.^23–25^ NPS-detected viral loads in patients with covid-19 rapidly decrease in the upper airways in those with increasing disease severity as the viral load shifts to the lower respiratory tract.^23–25^ We have previously detected surfactant protein A in FMS samples, indicating the likelihood that FMS captures at least some of its content from the lower respiratory tract.^26^ This feature provides a potential explanation for why higher FMS viral loads detected here correlated with higher ISARIC scores and therefore predicted higher disease severity in contrast to NPS which does not. Whilst larger, formally powered studies are needed, FMS may have a role in early identification of individuals at risk of deterioration, hospitalisation and death.

Our study was limited by size. As a pilot study using a novel device, we were unable to undertake formal power calculations. We assessed disease severity using ISARIC scoring systems rather than clinical outcomes of intubation or death, because our cohort had a small number of deaths. Ethnic minority groups, especially those from Asian ethnic groups are generally more likely to have worse outcomes from covid-19 compared to white groups^27,28^; the discordant findings here likely reflect of selection bias of younger Asian patients with less severe clinical disease. Our results should be verified in larger and more diverse groups of patients. Finally, the relationship of viral loads captured on FMS to replication-competent virus is uncertain and will be a key area for further research.

In conclusion, we present a novel, clinically compatible tool for the detection and quantification of exhaled SARS-CoV-2 in hospitalised patients with covid-19. We detected potentially significant relationships between exhaled viral load, clinical presentation and outcome. Our findings provide a strong incentive to investigate this approach further, to understand the pattern and quantity of exhaled virus both in hospitalised and community cases, and to explore the potential of FMS to identify those who are most infectious.

## Data Availability

Individual participant data will be available that underlie the results reported in this article after de-identification along with the study protocol between 9-36 months post publication.

## Acknowledgements

We gratefully acknowledge support from staff and patients of University Hospitals of Leicester in completing this work. The University of Leicester Core Biotechnology Services Containment Level 3 facility provided essential support. Ed Farries and colleagues at Qiagen are thanked for donated reagents and equipment enabling initiation of the work and we also thank Drs Julian Tang and David Jenkins for their advice.

## References

1 Meyerowitz EA, Richterman A, Ghandhi RT, Sax PE. Transmission of SARS-CoV-2: A review of viral, host and environmental factors. Ann Intern Med 2021;174:69–79.

2 Cevik Muge, Kuppalli Krutika, Kindrachuk Jason, Peiris Malik. Virology, transmission, and pathogenesis of SARS-CoV-2. BMJ 2020;371:1–6. Doi: 10.1136/bmj.m3862.

3 Lednicky John A., Lauzardo Michael, Fan Z. Hugh, Jutla Antarpreet S., Tilly Trevor B., Gangwar Mayank, et al. Viable SARS-CoV-2 in the air of a hospital room with COVID-19 patients. MedRxiv 2020;100:476–82. Doi: 10.1101/2020.08.03.20167395.

4 Lu Jianyun, Gu Jieni, Li Kuibiao, Xu Conghui, Su Wenzhe, Lai Zhisheng, et al. COVID-19 Outbreak associated with air conditioning in restaurant, Guangzhou, China, 2020. Emerg Infect Dis 2020;26(9):2298. Doi: 10.3201/eid2609.201749.

5 Ma Jianxin, Qi Xiao, Chen Haoxuan, Li Xinyue, Zhang Zheng, Wang Haibing, et al. Coronavirus Disease 2019 patients in earliar stages exhaled millions of severe acute respiratory syndrome coronavirus 2 per hour. Clin Infect Dis 2020;ciaa1283. Doi: https://doi.org/10.1093/cid/ciaa1283.

6 Asadi Sima, Cappa Christopher D., Barreda Santiago, Wexler Anthony S., Bouvier Nicole M., Ristenpart William D. Efficacy of masks and face coverings in controlling outward aerosol particle emission from expiratory activities. Sci Rep 2020;10(1):1–13. Doi: 10.1038/s41598-020-72798-7.

7 Fennelly Kevin P., Martyny John W., Fulton Kayte E., Orme Ian M., Cave Donald M., Heifets Leonid B. Cough-generated Aerosols of Mycobacterium tuberculosis: A New Method to Study Infectiousness. Am J Respir Crit Care Med 2004;169(5):604–9. Doi: 10.1164/rccm.200308-1101OC.

8 Huynh Kerrianne N., Oliver Brian G., Stelzer Sacha, Rawlinson William D., Tovey Euan R. A new method for sampling and detection of exhaled respiratory virus aerosols. Clin Infect Dis 2008;46(1):93–5. Doi: 10.1086/523000.

9 Kanaujia R., Biswal M., Angrup A., Ray P. Inhale, then exhale: start afresh to diagnose Severe Acute Respiratory Syndrome Coronavirus 2 (SARS-CoV-2) by non-invasive face-mask sampling technique. Clin Microbiol Infect 2020;26(12):1701–2. Doi: 10.1016/j.cmi.2020.06.034.

10 Kennedy Matthew, Ramsheh Mohammadali Y., Williams Caroline M.L., Auty Joss, Haldar Koirobi, Abdulwhhab Mohamad, et al. Face mask sampling reveals antimicrobial resistance genes in exhaled aerosols from patients with chronic obstructive pulmonary disease and healthy volunteers. BMJ Open Respir Res 2018;5(1):1–8. Doi: 10.1136/bmjresp-2018-000321.

11 Williams Caroline M.L., Cheah Eddy S.G., Malkin Joanne, Patel Hemu, Otu Jacob, Mlaga Kodjovi, et al. Face mask sampling for the detection of Mycobacterium tuberculosis in expelled aerosols. PLoS One 2014;9(8):1–7. Doi: 10.1371/journal.pone.0104921.

12 Williams Caroline M., Abdulwhhab Mohamad, Birring Surinder S., De Kock Elsabe, Garton Natalie J., Townsend Eleanor, et al. Exhaled Mycobacterium tuberculosis output and detection of subclinical disease by face-mask sampling: prospective observational studies. Lancet Infect Dis 2020;20(5):607–17. Doi: 10.1016/S1473-3099(19)30707-8.

13 Knight Stephen R., Ho Antonia, Pius Riinu, Buchan Iain, Carson Gail, Drake Thomas M., et al. Risk stratification of patients admitted to hospital with covid-19 using the ISARIC WHO Clinical Characterisation Protocol: Development and validation of the 4C Mortality Score. BMJ 2020;370(September):1–13. Doi: 10.1136/bmj.m3339.

14 Gupta Rishi K, Harrison Ewen M, Ho Antonia, Docherty Annemarie B, Knight Stephen R, van Smeden Maarten, et al. Development and validation of the ISARIC 4C Deterioration model for adults hospitalised with COVID-19: a prospective cohort study. Lancet Respir Med 2021;2600(20). Doi: 10.1016/s2213-2600(20)30559-2.

15 Al-Taie Alaa, Han Xiaoxiao, Williams Caroline M., Abdulwhhab Mohamad, Abbott Andrew P., Goddard Alex, et al. 3-D printed polyvinyl alcohol matrix for detection of airborne pathogens in respiratory bacterial infections. Microbiol Res 2020;241(August):126587. Doi: 10.1016/j.micres.2020.126587.

16 Han Mi Seon, Byun Jung Hyun, Cho Yonggeun, Rim John Hoon. RT-PCR for SARS-CoV-2: quantitative versus qualitative. Lancet Infect Dis 2020;21(2):165. Doi: 10.1016/S1473-3099(20)30424-2.

17 Sriraman Kalpana, Shaikh Ambreen, Parikh Swapneil, Udupa Shreevatsa, Chatterjee Nirjhar, Shastri Jayanthi, et al. Non-Invasive Sampling Using an Adapted N-95 Mask: An Alternative Method to Quantify SARS-CoV-2 in Expelled Respiratory Samples and Its Implications in Transmission. SSRN Electron J 2020. Doi: 10.2139/ssrn.3725611.

18 Cevik Muge, Tate Matthew, Lloyd Ollie, Maraolo Alberto Enrico, Schafers Jenna, Ho Antonia. SARS-CoV-2, SARS-CoV, and MERS-CoV viral load dynamics, duration of viral shedding, and infectiousness: a systematic review and meta-analysis. The Lancet Microbe 2020;5247(20):1–10. Doi: 10.1016/s2666-5247(20)30172-5.

19 Sze Shirley, Pan Daniel, Williams Caroline ML, Barker Joseph, Minhas Jatinder S, Miller Chris J, et al. The need for improved discharge criteria for hospitalised patients with COVID-19—implications for patients in long-term care facilities. Age Ageing 2020. Doi: 10.1093/ageing/afaa206.

20 Teranaka Wakana, Pan Daniel. Discharge criteria for patients with COVID-19 to long-term care facilities requires modification. Clin Med (Northfield Il) 2021;21(1):e116.2-e117. Doi: 10.7861/clinmed.let.21.1.2.

21 Cui Chunguang, Shin Kyeong-Ryeol, Bae Joon-Yong, Kweon Oh-Joo, Lee Mi-Kyung, Choi Seong-Ho, et al. Duration of culturable SARS-CoV-2 in hospitalised patients with covid-19. N Engl J Med 2021;2020:2020–2. Doi: 10.1056/nejmc2026670.

22 Mina Michael J, Peto Tim E, Garcia-Finana Marta, Semple Malcolm G, Buchan Iain E. Clarifying the evidence on SARS-CoV-2 antigen rapid tests in public health responses to COVID-19 2021;6736(21):21–3. Doi: 10.1016/S0140-6736(21)00425-6.

23 Wölfel Roman, Corman Victor M., Guggemos Wolfgang, Seilmaier Michael, Zange Sabine, Müller Marcel A., et al. Virological assessment of hospitalized patients with COVID-2019. Nature 2020;581(7809):465–9. Doi: 10.1038/s41586-020-2196-x.

24 To Kelvin Kai Wang, Tsang Owen Tak Yin, Leung Wai Shing, Tam Anthony Raymond, Wu Tak Chiu, Lung David Christopher, et al. Temporal profiles of viral load in posterior oropharyngeal saliva samples and serum antibody responses during infection by SARS-CoV-2: an observational cohort study. Lancet Infect Dis 2020;20(5):565–74. Doi: 10.1016/S1473-3099(20)30196-1.

25 Liu Yang, Yan Li Meng, Wan Lagen, Xiang Tian Xin, Le Aiping, Liu Jia Ming, et al. Viral dynamics in mild and severe cases of COVID-19. Lancet Infect Dis 2020;20(6):656–7. Doi: 10.1016/S1473-3099(20)30232-2.

26 Abdhulwhhab Mohamad T. Use of Face-Mask Sampling as a Means of Characterising the Microbiota Exhaled from Human Respiratory Tract in Health and Disease Thesis submitted for the degree of Doctor of Philosophy at the University of Leicester by Dr Mohamad Tayser Abdulwhhab, MD (. 2019.

27 Sze S, D Pan, Nevill CR, Gray LJ, Martin CA, Nazareth J, et al. Ethnicity and clinical outcomes in COVID-19: A systematic review and meta-analysis. EClinicalMedicine 2020. Doi: 10.1016/j.eclinm.2020.100630.

28 Pan Daniel, Sze Shirley, Martin CA, Nevill CR, Minhas JS, Divall P, et al. The missing link in ethnicity and covid-19 research—time to separate the risk of infection from the risk of severe disease. BMJ Blogs. Available at https://blogs.bmj.com/bmj/2021/01/29/the-missing-link-in-ethnicity-and-covid-19-research-time-to-separate-the-risk-of-infection-from-the-risk-of-severe-disease/. Accessed February 9, 2021, 2021.

