## Supplementary files for "Exhaled SARS-CoV-2 quantified by face-mask sampling in hospitalised patients with covid-19"

and
University Hospitals of Leicester NHS Trust, United Kingdom

### Figure S1: Duckbill mask with 4 PVA strips

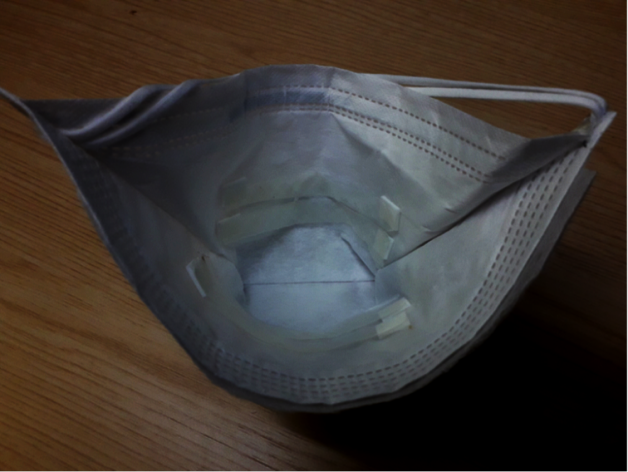

### Detailed Methods

Mask Strip Processing: Following exposure, masks were returned to their labelled plastic transport bags (double layered) taking care not to bring exposed PVA strips in contact with each other or increasing contact with mask material until they had dried out. Sealed samples were maintained at ambient temperature and processed within seven days of sampling.

For processing, two PVA strips were aseptically cut from the sampling mask and placed within a 15 ml polypropylene centrifuge tube (Corning, US). For dissolution, the following were added to the strips: 4 ml ultrapure molecular grade water and, from a QIAamp DSP Circulating Nucleic Acid Kit (Qiagen, Gernamy, cat no.61504), 3.2 ml ACL buffer and 400 µl Proteinase K and 1.12 µg carrier RNA (cRNA), together with 50 µl of our internal control virus (HCoV-OC43, see below). Following vortexing (30 s), the sample was treated at 70⁰C in a water bath for 10 min. RNA was then extracted from the supernatant using the Qiagen kit as above but omitting the 60⁰C and 4⁰C incubation steps. RNA was eluted in 45 µl of buffer AVE and stored at -80⁰C.

Swab processing: Combined nose and throat swabs were collected using cotton swabs (Deltalab, Spain). Immediately after collection the swab head was removed and stored dry in a 2ml vial (Sarstedt, UK). Adsorbed sample was released from the swab by the addition of 500 µl mH_2_O followed by vortexing for 15 s. 100 µl of swab eluate was added to 350 µl RLT buffer (RNeasy Mini kit; Qiagen, Germany cat no. 74104) containing 1.12 µg cRNA (QIAamp DSP Circulating Nucleic Acid Kit Qiagen, Germany cat no. 61504). RNA was then extracted using the RNeasy mini kit according to the manufacturer’s instructions.

Internal and positive controls: Human Coronavirus HCoV-OC43 was prepared from clarified (500 x g, 10 min) 5 day Vero E6 infection supernatants (virus isolate and cells kindly provided respectively by Professor Kin-Chow Chang and Dr Richard Urbanowicz, University of Nottingham) and frozen at -80⁰C. Aliquots of dilutions were prepared to yield approximate Cts of ~32 (OC43 RT-PCR) when added to samples.

For SARS-CoV-2 positive controls, one extracted with each run of patient samples, dilutions of a residual positive diagnostic sample were prepared to give Cts of ~23 by E assay after addition to PVA strips, heating to 70°C for 10 min and dissolving (as described above for patient sample strips) and storage at -80⁰C until required to be added to each batch of RNA extraction and assay run.

For *in vitro* studies SARS-CoV2 was isolated in Vero E6 cell culture by limiting dilution of a RT-qPCR positive swab sample; confirmation of identity at passage 3 was undertaken by E and RdRp-Hel assay ^1, 2^.

RT-qPCR: RNA was detected and quantified using the QuantiNova Probe RT-PCR Kit (Qiagen, Cat: 208356) and a Rotor-Gene Q thermocyler (Qiagen, Cat 9001590). Mastermix composition, primers and probes are shown in Table S1. Cycling conditions were the same for all three targets: 45⁰C, 10 min; 95⁰C, 5 min; and 95⁰C, 5 s followed by 60⁰C, 30 s for 45 cycles. Assays runs were accepted if the internal and positive control values fell within 2 Cts of the mean.

### Table S1 Assay composition

| Target | | E (Sarbeco)^1^ | | RdRp-Hel^2^ | | OC43^3^ |
| --- | --- | --- | --- | --- | --- | --- |
| Forward | | ACAGGTACGTTAATAGTTAATAGCGT | | CGCATACAGTCTTRCAGGCT | | ATGTTAGGCCGATAATTGAGGACTAT |
| Reverse | | ATATTGCAGCAGTACGCACACA | | TGTGATGTTGAWATGACATGGTC | | AATGTAAAGATGGCCGCGTATT |
| Probe | | ACACTAGCCATCCTTACTGCGCTTCG [5']Fam [3']BHQ-1 | | TTAAGATGTGGTGCTTGCATACGTAGAC [5']Fam [3']IABkFQ | | CATACTCTGACGGTCACA AT [5’] HEX [3’]IABkFQ |
| MASTERMIX | | | | | | |
| Primers (10mM) | | 1.6 µl, (final 800 nM) | | 2 µl, (final 800 nM) | | 1.6 µl, (final 800 nM) |
| Probe (10M) | | 0.4 µl, (final 200 nM) | | 0.5 µl, (final 200 nM) | | 0.4 µl, (final 100 nM) |
| 2x reaction mix | | 10 µl | | 12.5 µl | | 10 µl |
| RT Probe Mix | | 0.2 µl | | 0.25 µl | | 0.2 µl |
| mH_2_O | | 1.2 µl | | 2.75 µl | | 1.2 µl |
| Template | | 5 µl | | 5 µl | | 5 µl |
| Total | | 20 µl | | 25 µl | | 20 µl |

Assay Performance: All assays included positive and negative (no template) controls. Positive controls were included prior to extraction and comprised viral transport medium from a strongly-positive patient swab sample diluted in mH_2_O. For mask assays this was added to dissolved PVA in aliquots equivalent to two dissolved strips then stored at -80⁰C. Assay runs were accepted if the positive control fell within 1 Ct of the established mean value. The OC43 internal control was added to all dissolved samples prior to extraction. Individual assays were accepted if the related OC43 result was within 2 Cts of the established mean (n=74). All runs revealed negative sample results and all negative controls gave no Ct endpoints. The technical performance of the RT-qPCRs on extracted RNA was compared to the local NHS lab and showed 95% correspondence with positive/negative designation (20 RNA extracts diluted 1:3) and agreement in the ranking of signal strengths.

Preservation of signal strength on storage: This was tested by directly spiking PVA strips in triplicate with our SARS-CoV-2 isolate and storing dry at room temperature for two and seven days. No difference in the baseline Ct of 15.8 was discernible at 2 days and a decline of less than one Ct was seen at seven days (16.7). In addition, we assayed spare strips from a second location within face mask samples taken early in our studies and compared signals in the stored RNA with those obtained from a new extraction. Table S2 shows that there was no loss of signal positivity during storage. Indeed, some samples revealed substantially lower Cts. While these results give confidence that signals are not lost on storage they also indicate that strips placed at different locations in the mask may give different results

|  | **RdRp-Hel Ct Initial extract** | **RdRp-Hel Ct ~3 months** |
| --- | --- | --- |
| A12 | 30.85 | 30.5 |
| A13 | 23.95 | 20.2 |
| A16 | 34.8 | 34.42 |
| A20 | 36.4 | 29.94 |
| A22 | 30.43 | 27.12 |
| B01 | 32.12 | 32.3 |
| B06 | 28.37 | 39.16 |

Table S2 Effect of strip storage for ~3 months
Comparison of Cts from RNA extracts obtained and stored within 72h of sampling and those from PVA strips stored for ~3 months prior to extraction. Early assays were performed with RdRp-Hel primers and these were used again here for compatibility.

### Quantitation Standards

**Preparation:** Synthetic Sars-CoV-2 RNA (Wuhuhan-Hu-1/MN908947.3, Twist Bioscience ,USA) was diluted in AVE buffer containing cRNA to produce standards from 2x10^1^ – 2x10^4^ copies/µl. 5µl of each standard was assayed in triplicate using the E RT-qPCR assay to produce a standard curve with a dynamic range of 10^2^-10^5^ copies per reaction.

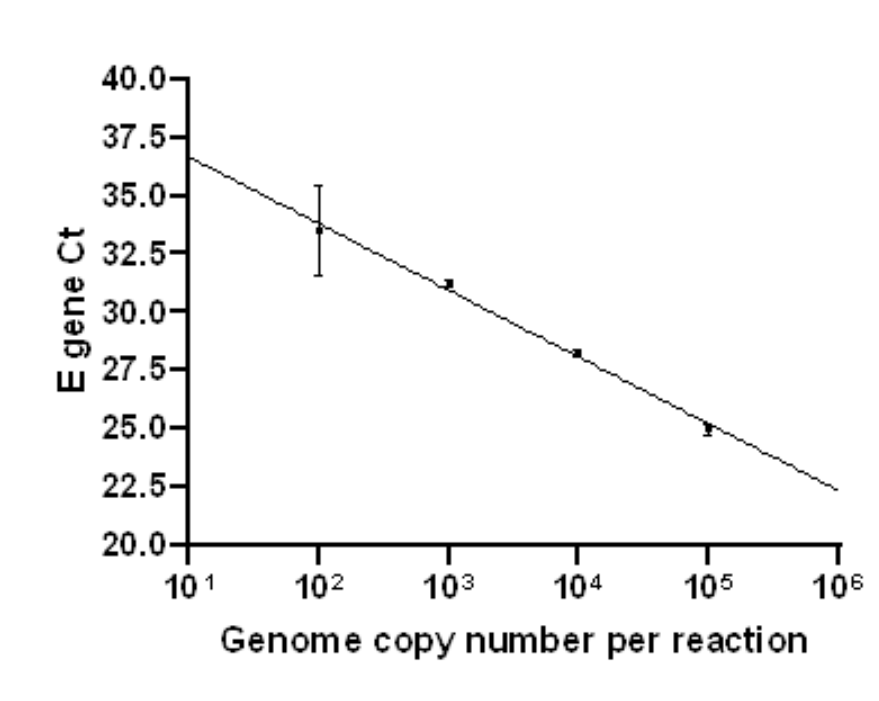

**Assignment of values:** Cts falling within the range above were assigned assay values from the standard curve. A scaling factor of 8 was applied to give genome copies per strip and 16 to give copies per 100 µl swab eluate. Positive assays with Cts outside this range (repeat positive or second target (RdRp) positive) were assigned values of 8 or 16 for FMS and NPS respectively.

### Figure S2: Frequency distributions of FMS and NPS results

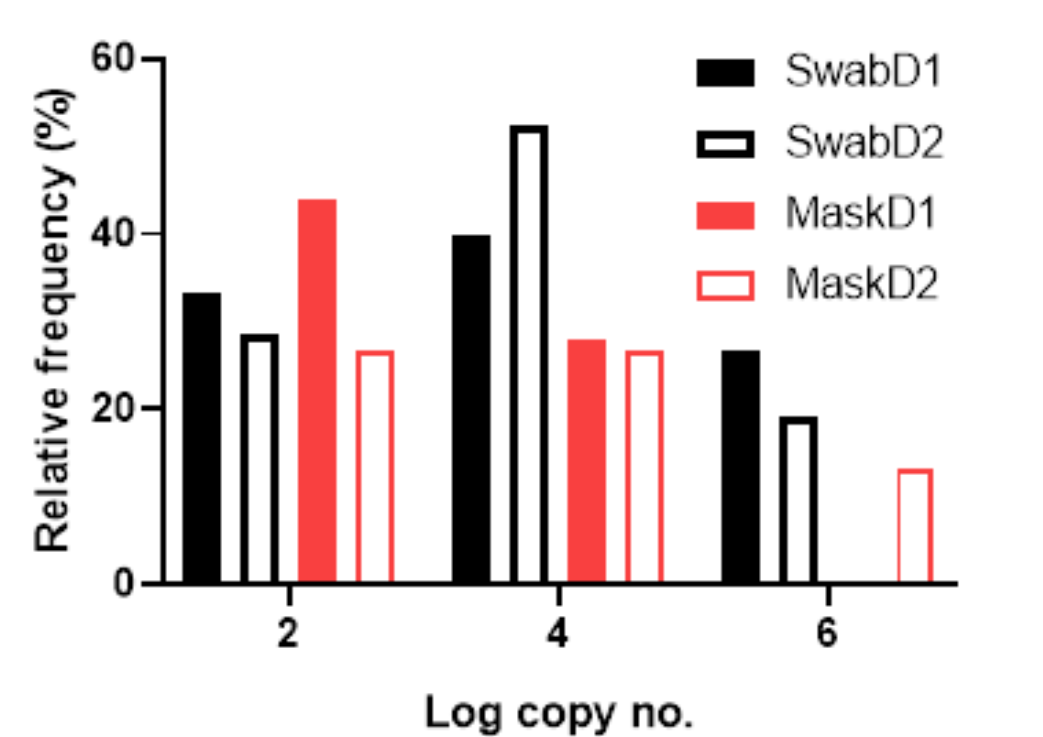

**SwabD1 – NPS day 1; SwabD2 – NPS day 2; MaskD1 – FMS day 1; MaskD2 – FMS day 2**

### References

1. Corman, V. M., O. Landt, M. Kaiser, R. Molenkamp, A. Meijer, D. K. W. Chu, T. Bleicker, S. Brunink, J. Schneider, M. L. Schmidt, D. Mulders, B. L. Haagmans, B. van der Veer, S. van den Brink, L. Wijsman, G. Goderski, J. L. Romette, J. Ellis, M. Zambon, M. Peiris, H. Goossens, C. Reusken, M. P. G. Koopmans and C. Drosten (2020). "Detection of 2019 novel coronavirus (2019-nCoV) by real-time RT-PCR." Eurosurveillance **25**(3): 23-30.
2. Chan, J. F., C. C. Yip, K. K. To, T. H. Tang, S. C. Wong, K. H. Leung, A. Y. Fung, A. C. Ng, Z. Zou, H. W. Tsoi, G. K. Choi, A. R. Tam, V. C. Cheng, K. H. Chan, O. T. Tsang and K. Y. Yuen (2020). "Improved Molecular Diagnosis of COVID-19 by the Novel, Highly Sensitive and Specific COVID-19-RdRp/Hel Real-Time Reverse Transcription-PCR Assay Validated In Vitro and with Clinical Specimens." J Clin Microbiol **58**(5): e00310-20.
3. Vijgen, L., E. Keyaerts, E. Moes, P. Maes, G. Duson and M. Van Ranst (2005). "Development of one-step, real-time, quantitative reverse transcriptase PCR assays for absolute quantitation of human coronaviruses OC43 and 229E." J Clin Microbiol **43**(11): 5452-5456.
